# Accuracy and reliability of data extraction for systematic reviews using large language models: A protocol for a prospective study

**DOI:** 10.1101/2024.05.22.24307740

**Authors:** Takehiko Oami, Yohei Okada, Taka-aki Nakada

## Abstract

**Background:** Systematic reviews require extensive time and effort to manually extract and synthesize data from numerous screened studies. This study aims to investigate the ability of large language models (LLMs) to automate data extraction with high accuracy and minimal bias, using clinical questions (CQs) of the Japanese Clinical Practice Guidelines for Management of Sepsis and Septic Shock (J-SSCG) 2024. the study will evaluate the accuracy of three LLMs and optimize their command prompts to enhance accuracy.

**Methods:** This prospective study will objectively evaluate the accuracy and reliability of the extracted data from selected literature in the systematic review process in J-SSCG 2024 using three LLMs (GPT-4 Turbo, Claude 3, and Gemini 1.5 Pro). Detailed assessment of errors will be determined according to the predefined criteria for further improvement. Additionally, the time to complete each task will be measured and compared among the three LLMs. Following the primary analysis, we will optimize the original command with integration of prompt engineering techniques in the secondary analysis.

**Trial registration:** This research is submitted with the University hospital medical information network clinical trial registry (UMIN-CTR) [UMIN000054461].

**Conflicts of interest:** All authors declare no conflicts of interest to have.

## Background

Systematic reviews are essential for synthesizing research findings and providing reliable evidence for decision-making in healthcare settings. Traditionally, these reviews have been labor-intensive, requiring extensive time and effort to manually extract and synthesize data from numerous studies [1-3]. The advent of advanced natural language processing and artificial intelligence (AI) technologies offers promising avenues to enhance the efficiency and accuracy of systematic reviews [4-6].

Recent advancements have shown that AI can effectively support data extraction by acting as a second reviewer, enhancing both the speed and reliability of data extraction from structured and semi-structured research papers [7-9]. Prior studies, including the recent proof-of-concept utilizing an LLM, have demonstrated promising results in automating data extraction processes, achieving high accuracy and reliability while substantially reducing human error and labor intensity [10-12]. Despite these advancements, the potential of LLMs in handling diverse and complex datasets across broader domains of medical research remains underexplored. In addition, the performance of different LLMs for data extraction has yet to be fully explored.

This study aims to explore and evaluate the effectiveness of LLMs in systematic reviews, focusing on their potential to automate data extraction while ensuring high accuracy and minimal bias. Using the clinical questions (CQs) from the Japanese Clinical Practice Guidelines for the Management of Sepsis and Septic Shock (J-SSCG), we will assess the accuracy of two LLMs and optimize the command prompt to achieve higher accuracy.

## Methods

### Study design and settings

We will conduct a prospective study to evaluate the accuracy and reliability of LLMs for data extraction in systematic reviews. This research will involve several phases: the elaboration of a suitable query for LLMs based on the prompt engineering technique, an automated data extraction process, and evaluate the accuracy of the extracted data. To maintain transparency and allow for reliability, our review protocol has been recorded and shared via the medRxiv pre-print server. Furthermore, our research is officially listed in the University Hospital Medical Information Network (UMIN) clinical trials registry, with the registration number [UMIN000054461]. This registration highlights our dedication to maintaining high scientific and ethical standards in our research.

### Japanese Clinical Practice Guidelines for the Management of Sepsis and Septic Shock

The efficiency of LLMs will be assessed using the clinical questions (CQs) from the J-SSCG 2024, as outlined in our earlier report [5]. This guideline was created by the Japanese Society of Intensive Care Medicine (JSICM) and the Japanese Association for Acute Medicine (JAAM), following the J-SSCG 2020 and addressing unique clinical scenarios in Japan related to sepsis and septic shock [13].

The same five CQs (see Table 1) utilized in our previous analysis will be employed in this study [5]. Comprehensive literature searches were conducted across CENTRAL, PubMed, and Ichushi-Web for these CQs, with the working group members developing an exhaustive search strategy to encompass all pertinent studies. The literature review was restricted to works published in Japanese and English. All relevant titles and abstracts were systematically downloaded, organized, and duplicates were removed using EndNote, which serves as the reference management tool for J-SSCG 2024. After title/abstract and full-text screening, the final set of literature was selected for the qualitative analysis. The J-SSCG 2024 members extracted background information and outcomes in each study. In this study, we will use the selected publications from the five CQs for assessing the accuracy of the data extraction.

**Table 1.** The list of the patient/population/problem, intervention, and comparison of the selected clinical questions.

|  | Patient, population, problem | Intervention | Comparison |
| --- | --- | --- | --- |
| CQ1 | Adult patients (18 years old or older) diagnosed with or suspected of having infection, bacteremia, or sepsis | Balanced crystalloid administration | 0.9% sodium chloride administration |
| CQ2 | Adult patients (18 years old or older) with sepsis, or suspected as sepsis, infection, bacteremia or patients admitted to ICU | Targeting a higher mean arterial pressure | Targeting a lower mean arterial pressure |
| CQ3 | Adult patients (18 years old or older) with sepsis presenting with severe metabolic acidosis or patients admitted to ICU | Sodium bicarbonate administration | No sodium bicarbonate administration |
| CQ4 | Adult patients (18 years old or older) with sepsis or septic shock | Usual care with at least one of the following tissue perfusion parameters: lactate/lactate clearance, capillary refill time, ScvO<sub>2</sub>/SvO<sub>2</sub>, and P(v-a) CO<sub>2</sub>/C (a-v) O<sub>2</sub>. | Usual care with different parameters mentioned in the interventional group, or standard care without the utilization of any specific tissue perfusion parameters |
| CQ5 | Adult patients (18 years old or older) with sepsis, sepsis-induced hypotension, or septic shock | Restrictive fluid management, which aims to reduce the amount of fluid therapy for up to 24 h | Conventional fluid management or non-restrictive fluid management defined by authors |

CQ: clinical question; ICU: intensive care unit

### Large language model

We will use following three LLM; GPT-4 Turbo (OpenAI, San Francisco, CA, USA), released on November 7, 2023, Claude 3 (Anthropic, San Francisco, CA, USA), released on March 14, 2024, and Gemini 1.5 Pro (Google DeepMind, Mountain View, CA, USA), released on April 21, 2024, to investigate the feasibility, accuracy and reliability for data extraction in systematic reviews. If these LLMs are updated or other novel LLM become available, and it is suitable for this study aim, we will include it for evaluation. These LLMs can recognize portable document format (PDF) files and export the output based on the user’s request. After importing the PDF files of the final set of literature for the qualitative analysis on our folders, we will upload the PDF files on the LLMs through a user interface and implement the automated process of data extraction using the following command prompts. We submitted a request to the responsible developers or ensured a privacy policy to exclude the information that users input into LLMs from the training data. Prior to this process, unreadable PDF files were converted into readable ones using the optical character recognition feature of Adobe Acrobat (Adobe Inc., San Jose, CA, USA). Of note, we will divide the data extraction processes into two parts: background information of the studies and outcomes. If PDF file is not suitable for optical character recognition such as the file is based on scanning of a paper article and the quality of document is inappropriate, we will exclude the PDF for evaluation.

Prompt for extracting background information:

We will use the following prompt to extract the background information.

“You are conducting a systematic review and meta-analysis, focusing on a specific area of medical research. Your task is to read through the imported PDF file and to extract background information about study and participants. Please summarize the extracted information and export it to an Excel file. If there is no relevant information in the PDF file, please enter “not available” in the cell. If the word count of the extracted data for each information is over 300 words, please summarize the descriptions and indicate that the content is being summarized. The required information is as follows:

\# Title of the article
\# Name of first author
\# Year of publication
\# Journal name
\# Study design
\# Inclusion criteria
\# Exclusion criteria
\# Number of participants to be randomized
\# Number of participants enrolled in the intervention and control groups
\# Average age of study participants in the intervention and control groups
\# Sex of participants in the intervention and control groups
\# Severity scores or scales such as Sequential Organ Failure Assessment (SOFA) and Acute Physiology and Chronic Health Evaluation (APACHE) in the intervention group and the control groups
\# Details of interventions
\# Details of controls

Prompt for outcome extraction:

We will use the following prompt to extract the background information.

“You are conducting a systematic review and meta-analysis, focusing on a specific area of medical research. Your task is to read through the imported PDF file and to extract outcome information about the participants. In extracting the outcomes, outcomes based on the intention-to-treat (ITT) analysis should be prioritized if available. If there is no outcome data based on the ITT analysis, please extract outcomes based on the per protocol analysis. If there is no clear description of the type of analysis written, please use the results from the main analysis. In such cases, please add the type of analysis, including ITT, per protocol, or unknown to the extracted data. Please summarize the extracted information and export it to an Excel file.

If the outcome is a dichotomous variable, please extract the number of events and participants in each group. If the outcome is a continuous variable, please extract the mean and standard deviation with the number of participants. If the outcome is provided with median and interquartile range (IQR) or mean and 95% confidence interval, please extract them as they are and add the description what the number is. If actual values are not provided in the text or table, please estimate the outcome value from figures or tables in the PDF file. Please add the type of continuous variables according to the extracted subjects. The list of outcome variables is as follows:

\# Outcome variable 1
\# Outcome variable 2
\# Outcome variable 3
\# Outcome variable 4”

### Data collection

Our research will evaluate the accuracy of the extracted data by the LLMs setting the reference standard is the results extracted by two or more human reviewers with the conventional method of systematic reviews. This conventional data extraction was conducted by J-SSCG 2024 members. If we find any error or missing in the human-led extracted data for reference standard in the research process, we will revise the reference standard by referring the original manuscript, following guidance by the Agency for Healthcare Research and Quality [14]. The extracted background information and outcomes will be used as standard references in this study to evaluate the accuracy. Two independent reviewers will evaluate the accuracy of the extracted data using the LLMs. If the reviewers will not reach an agreement, the third reviewer will resolve the conflict.

We will assess the error according to the original criteria (Table 2). Depending on the extent of accuracy in the extracted data on background and impact on the meta-analysis due to the errors in extracted data on outcomes, we will categorize the error of the extracted data into major errors, minor errors, and no errors. Additionally, we will categorize the error into the four types of errors: missing data, incorrect data, fabricated data, and others. If contents or errors which are not suitable for evaluation by two reviewers, we will not assess them and record the reason for the exclusion.

**Table 2.** Assessment of accuracy in data extraction.

|  | Background information | Outcome |
| --- | --- | --- |
| Major errors | Significant issues which can influence the credibility or results of systematic review and meta-analysis, such as more than half of the extracted information are missed, or any amount of important information are incorrect, missed, or fabricated. | Issues that can change the results of SR or meta-analysis, potentially leading to incorrect conclusions. E.g., incorrect number of randomized patients, incorrect number of patients assigned to intervention or control groups, incorrect number of events in outcome measures, or incorrect items. |
| Minor errors | Approximately most of the information (60% or more) is correctly extracted based on the reference standard, but some minor issues happen, such as some part of the information (20-40%) is not extracted even though it is shown in the paper, or inaccurately extracted. But errors are unlikely to influence the SR and meta-analysis. | Minor issues which are unlikely to change the results of SR or meta-analysis or lead to incorrect conclusions. E.g., errors in rounding. |
| No errors | Approximately 80% or more of the information is correctly and comprehensively extracted based on the reference standard. | No issues with the numbers. |

SR: systematic review

**Table 3.**
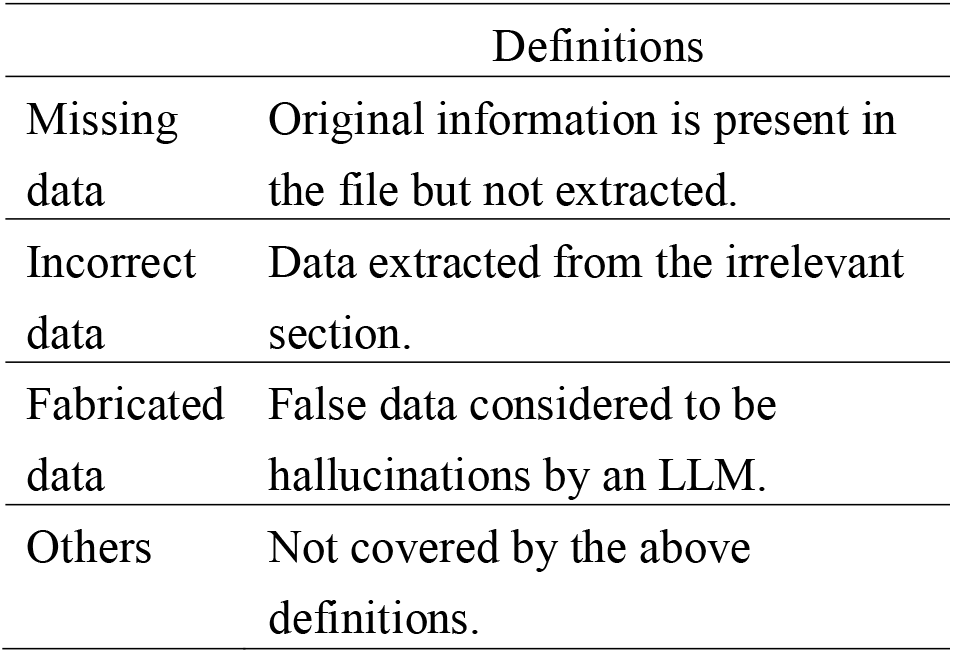
Type of errors in data extraction.

To ensure the reliability, we will repeat the same processes two times within two or six weeks after the first data extraction session. We will also measure the processing time to complete each data extraction session.

### Analysis

As a primary analysis, this study will make the confusion table and investigate the accuracy of the LLM performance in data extraction tasks using the original prompt without modification. The accuracy will be evaluated setting the results of the systematic reviews conducted in the process of J-SSCG as the reference standard. In the secondary analysis, we will integrate some modification (the chain-of-thought strategy, multiple voting strategy, and self-correction strategy) into the original prompt and evaluate the accuracy of the LLMs for data extraction [15, 16]. Furthermore, we will conduct the following additional analysis to understand the performance of LLM. (1) To assess the reliability, we will evaluate the results from second and third session as well as the first session and assess the accordance with the first session. Specifically, we will categorize them into matches (both correct or both incorrect) and discrepancies (one correct or one incorrect), to determine the extent of consistency in success or failure. (2) We will summarize and describe the characteristics of errors to understand the potential mechanism of errors. (3) We will summarize the time of LLM data extraction and compare it between different models if appropriate.

Continuous variables will be presented as means with standard deviations or medians with interquartile ranges, depending on their distribution. Categorical variables will be expressed as absolute numbers and percentages as appropriate. Comparisons among multiple groups were conducted using one-way ANOVA with subsequent Tukey posttest or Kruskal-Wallis test with Dunn’s multiple comparison test if required. For statistical analysis, GraphPad Prism 10 software (GraphPad Software, San Diego, CA, USA) will be utilized to manage research data and conduct statistical evaluations.

## Data Availability

All data produced in the present study are available upon reasonable request to the authors.

